# Development and validation of a length- and habitus-based method of total body weight estimation for critically ill adults requiring urgent weight-based medical intervention

**DOI:** 10.1101/2021.09.30.21264310

**Authors:** Mike Wells, Lara Nicole Goldstein, Giles Cattermole

**Affiliations:** Department of Emergency Medicine and Critical Care, Herbert Wertheim College of Medicine, Florida International University, Miami, FL, USA; Division of Emergency Medicine, Faculty of Health Sciences, University of the Witwatersrand, Johannesburg, South Africa; Emergency Department, Princess Royal University Hospital, King’s College Hospital NHS Trust, London, UK

## Abstract

**Background:** Erroneous weight estimation during the management of emergency presentations in adults may contribute to patient harm and poor outcomes. Patients can often not be weighed during emergencies and a weight estimation is required to facilitate weight-based therapies. Many existing methods of weight estimation are either unacceptably inaccurate or very difficult to use during the provision of emergency care.

**Methods:** The weight estimation system developed in this study was based on and modified from the PAWPER XL-MAC method, a paediatric weight estimation system that uses recumbent length and mid-arm circumference (MAC) to predict total body weight. This model was validated in the 2015 – 2018 National Health and Nutrition Examination Survey (NHANES) datasets. The primary outcome measure was to achieve >95% of estimations within 20% of measured weight (P20>95%).

**Results:** The modified PAWPER XL-MAC model achieved a P20 of 96.0% and a P10 of 71.3% in the validation dataset (N=11520). This accuracy (P20>95%) was maintained in both sexes, all ages, all ethnic groups, all lengths and in all habitus-types, except for the subgroup of severely obese individuals.

**Conclusions:** The modified PAWPER XL-MAC model proved to be a very accurate method of weight estimation. It is more accurate than most other published reports of existing methods of weight estimation, except for patients’ own estimations. It therefore could have a role in facilitating emergency drug dose calculations, if prospective studies bear out the accuracy found in this study.

## INTRODUCTION

There may be as much as a 250% increase in poor outcomes in stroke patients if their estimated weight, from which the dose of intravenous thrombolytics is calculated, differs from their actual total body weight by more than 10% [1]. This is important since as many as 85% of stroke patients may have thrombolytic doses calculated from estimated weights, when accurate measured weights cannot be obtained prior to initiating treatment [2, 3]. During medical emergencies it is frequently necessary to estimate weight to allow drug dose calculations, as obtaining a measured weight may not be possible or it may delay urgent treatment. If an estimated weight is inaccurate, it could give rise to critical medication dosing errors resulting in severe harm or even death [4]. The use of inaccurate methods of estimating weight, therefore, “cannot be considered to be good medical practice” [5]. Furthermore, it is essential that the weight estimation is as accurate as can be achieved without impacting negatively on resuscitative care by excessively consuming time or cognitive resources [6, 7]. This imperative for dosing accuracy applies not only to thrombolytic drugs, but many other emergently prescribed drugs including antimicrobials, anticoagulants, many cardiac medications, anticonvulsant, and antiepileptic medications [8].

Weight estimation in adults has not been as widely studied as in children, in whom the dual length- and habitus-based methods (such as the PAWPER XL tape and the Mercy method) have been established to be the most accurate [9-11]. In adults, the most accurate weight estimations generally come from the patients themselves, but patients are often not able to provide an estimation of their own weight, and self-estimates by overweight and obese patients are frequently inaccurate [12]. Other existing methods of weight estimation have not been shown to be sufficiently accurate for safe drug dosing and may be too complex or time-consuming to use during emergency care [3, 13].

Recently, attention has shifted to evaluating the best of the paediatric dual length- and habitus-based weight estimation systems in adults. The paediatric PAWPER XL-MAC method and the Mercy method have had a preliminary evaluation in adult populations, and outperformed other methods of weight estimation, except for patient self-estimations [14-16]. These methods have shown promise but would need to be modified and optimised for adults.

The selection of an important weight estimation method is important for both decision-makers and clinicians to develop appropriate policies and practices to ensure patient safety. Optimising dosing for weight-based drugs would maximise efficacy and minimise adverse effects

We hypothesized that the PAWPER XL-MAC method could be adapted for use in adults, to produce an accurate, easy-to-use weight estimation system. The aim of this study was, therefore, to develop and validate an adult version of the PAWPER XL-MAC tape, using recumbent length and mid-arm circumference (MAC) to predict total body weight.

## METHODS

This adult weight estimation model was based on the PAWPER XL-MAC tape paediatric weight estimation system [17]. The process used for developing, calibrating, and validating the new model is described below, with an overview in Figure 1.

**Figure 1.**
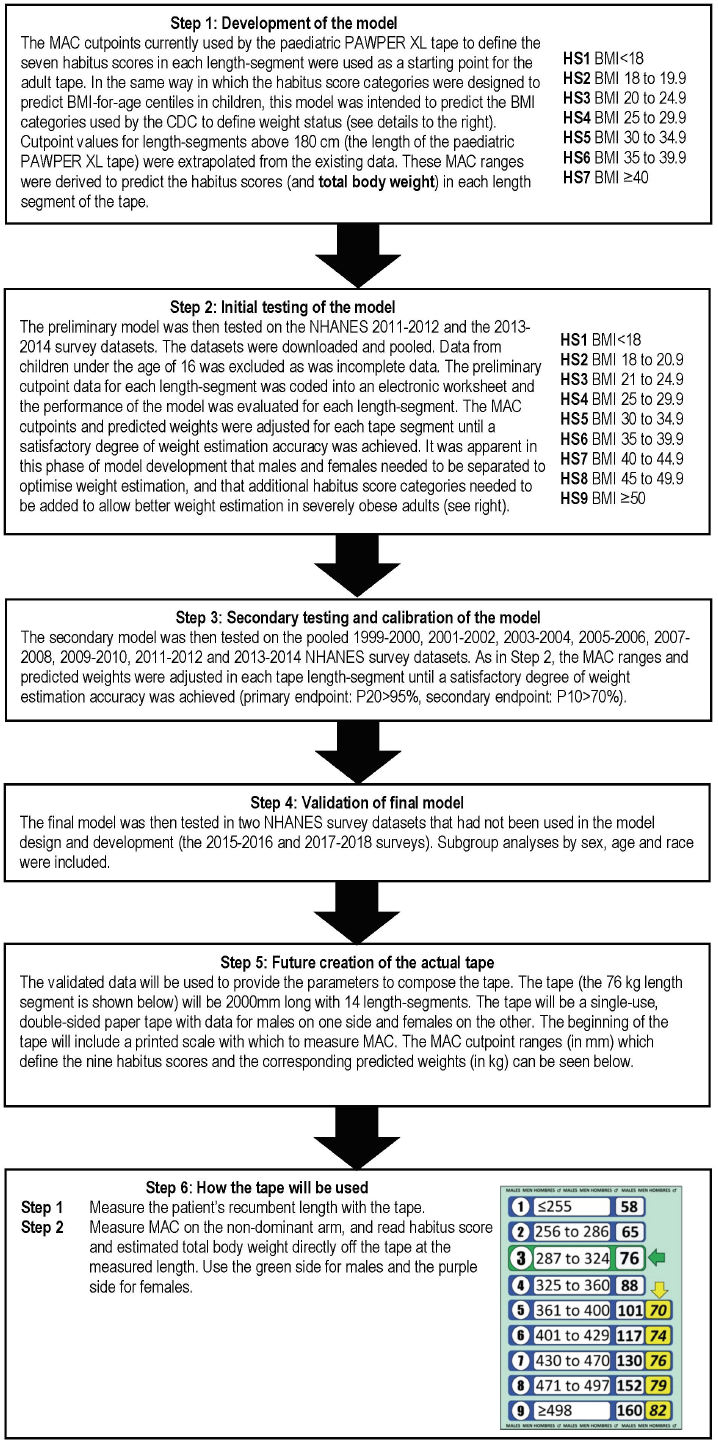
The methodology followed in this study. MAC = mid-arm circumference, BMI = body mass index, CDC = Centers for Disease Control and Prevention, HS = habitus score, NHANES = national health and nutrition examination survey, P10 = percentage of weight estimations within 10% of actual weight, P20 = percentage of weight estimations within 20% of actual weight,

### Method development

The PAWPER XL-MAC tape is used as follows: firstly, the tape is held alongside the patient, and the patient’s length measured from head to heel. The point where the patient’s heel crosses the tape is noted – this is the length-segment that provides the relevant weight-data. Secondly, MAC is measured from the patient’s non-dominant arm (ideally), using the scale on the PAWPER XL-MAC tape. From this measurement, the patient’s habitus score (HS) can be determined, and their predicted weight read directly off the tape. Each length-segment on the tape has MAC ranges to define each habitus score category. The habitus score categories range from HS1 to HS7, with HS1 representing a very underweight patient, HS3 a normal weight patient and HS7 a severely obese patient. The other habitus scores represent intermediate body types.

The existing length-segments, MAC cutpoint values and HS categories of the original paediatric PAWPER XL-MAC tape were used for the new provisional model. New length-segments were added to extend the length of the tape from 1800 mm to 2000 mm. The MAC cutpoints and predicted weights for these segments were extrapolated from data in the “shorter” segments. The model was then coded into an electronic worksheet (Microsoft Excel for Mac, 2020) and underwent a first stage of evaluation. The coding generated a weight estimation (kg) with an input of height (cm) and MAC (mm).

### Datasets

The National Health and Nutrition Examination Survey (NHANES) datasets from the 1999-2000 to the 2017-2018 surveys were downloaded from the Centers for Disease Control (CDC) website [18]. The downloaded data included the following variables: sequence number, sex, race, age, total body weight (TBW), height, body mass index (BMI) and MAC. Data from children under the age of 16 years was excluded, as were all individuals with incomplete data. The datasets were pooled as follows: the initial model testing was done in the NHANES 2011-2012 and 2013-2014 surveys, the model calibration was done in the NHANES 1999-2000, 2001-2002, 2003-2004, 2005-2006, 2007-2008, 2009-2010, 2011-2012 and 2013-2014 surveys, and the model validation was done in the 2015-2016 and 2017-2018 survey datasets.

### Initial model testing

The initial model was used to generate estimates of weight from height and MAC for each individual in the dataset. The data was then analysed separately for each length-segment of the tape (e.g., the 180.1 to 183.0 cm length segment). The specific outcomes that were then evaluated were: mean percentage error (MPE), which represented the estimation bias; the root mean square percentage error (RMSPE) which quantified the estimation precision; and the percentage of weight estimations that fell within 10% (P10) as well as within 20% (P20) of measured weight, which denoted overall accuracy.

After the initial testing it was apparent that the following changes needed to be made to the original model: males and females needed to be evaluated separately; two additional habitus score categories were added to accommodate weight estimations in severely obese adults; and the length-segments needed to be reduced in size. This new model was then calibrated in a larger pooled dataset.

### Model calibration

Weight estimates generated in the calibration dataset by the refined model were again examined in each length-segment of the tape. The MAC cutpoint values and predicted weights for each habitus score were adjusted until a target P20 >95% and P10 >70% was obtained for each habitus score category in each length-segment (based on previously suggested acceptable accuracy targets) [11]. At the upper limits of BMI there was a substantially reduced ability of MAC to discriminate between BMI categories. This suggested a biological limitation of using this single variable for further refinement of the model in these categories. The final model, shown in Table 1, was finally subjected to formal validation using unused data from the most recent NHANES surveys datasets.

**Table 1.**
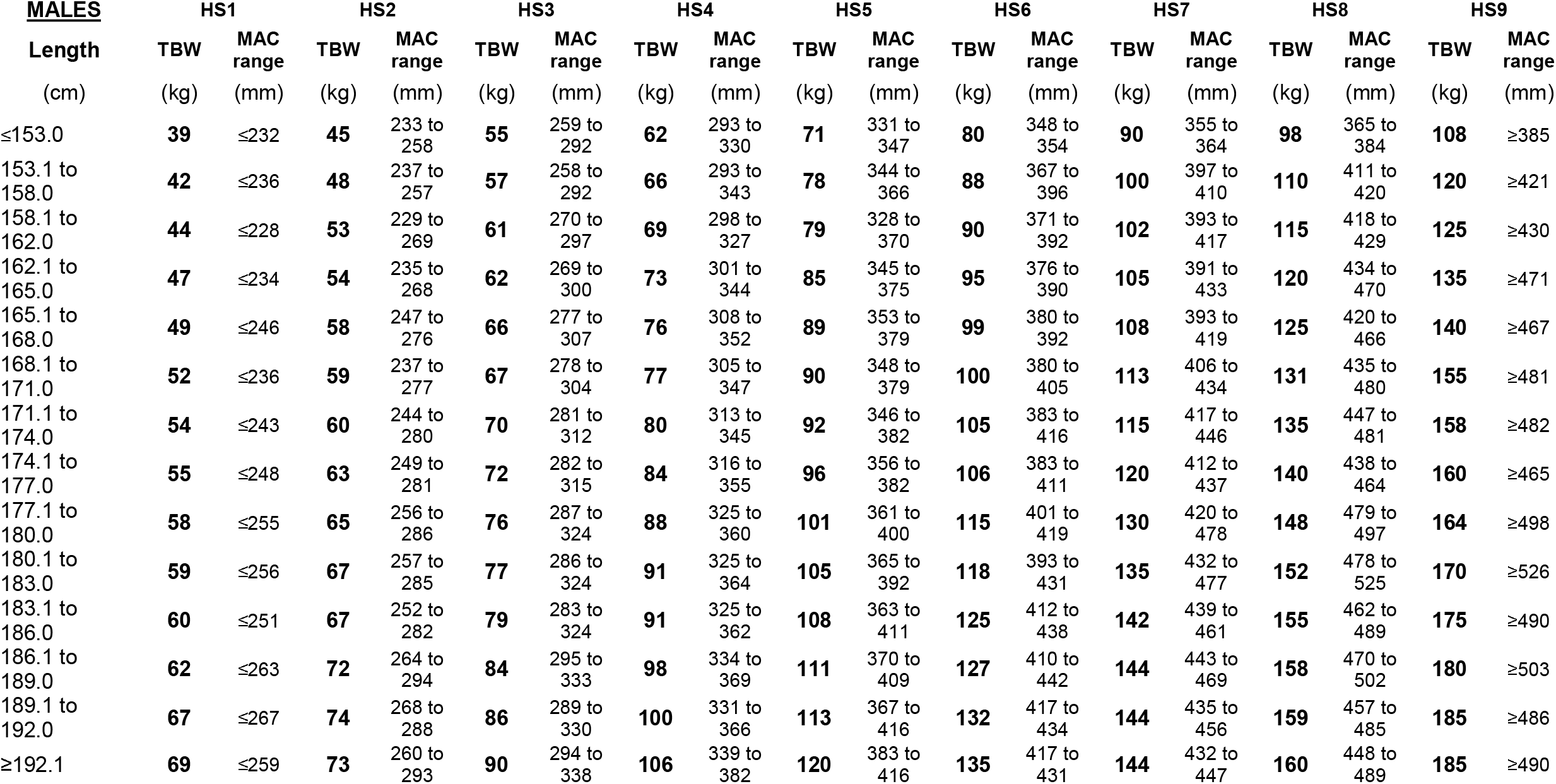
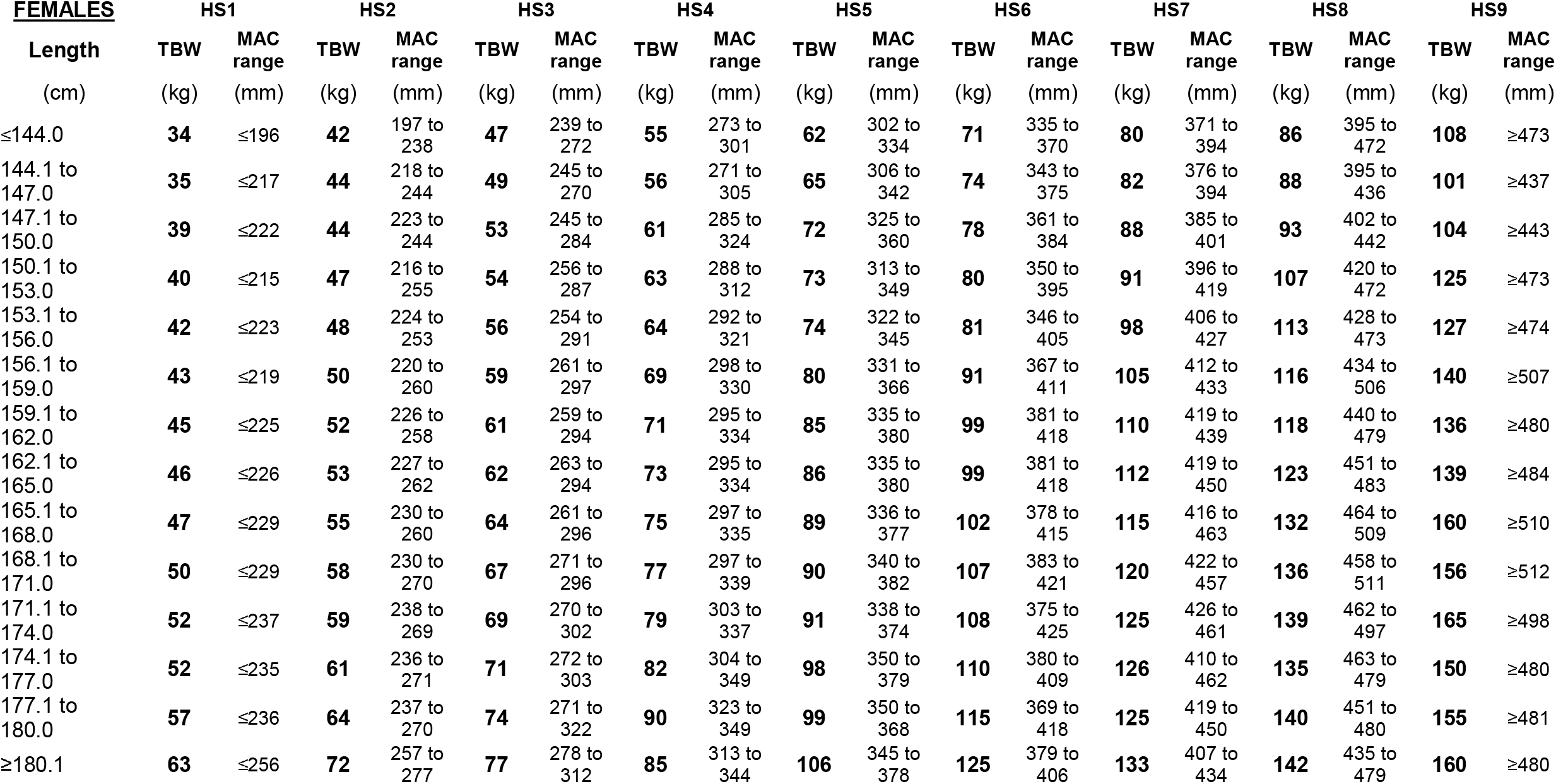
Final model for the male (upper panel) and female (lower panel) PAWPER XL-MAC adult tape. HS = habitus score, TBW = total body weight, MAC = mid-arm circumference.

### Model validation

The final model was validated by generating estimated weights in the pooled unused 2015-2016 and 2017-2018 NHANES survey datasets. The performance of the model was evaluated by comparing the estimated weights against actual measured weight using the MPE, RMSPE, P10 and P20. Subgroup analyses by sex, age, race (as defined in the NHANES datasets) and weight-status (determined by the CDC definitions of BMI as underweight, normal weight, overweight, obese, and severely obese) were performed. A Bland & Altman analysis was performed and graphically represented by subgroups of weight-status [19]. During the model development and analysis, the association between BMI and MAC was observed to be substantially weaker in severely obese patients than in thinner patients. For this reason, a correlation analysis between BMI and MAC was performed in these subgroups to evaluate and quantify this difference in the relationship.

All data was analysed using Microsoft Excel (Microsoft, Redmond, Washington: Microsoft, 2016) and Stata Statistical Software (StataCorp. 2019. Stata Statistical Software: Release 16. College Station, TX: StataCorp LLC). A significance level of 0.05 was used for all analyses.

### Outcome measures

The primary outcome measure was the performance of the model when compared to measured TBW. A P20 >95% and a P10 >70% was considered to be an acceptable accuracy of estimation, as has previously been suggested [11, 20]. This benchmark is regularly achieved in children by the paediatric dual length-based, habitus-modified weight-estimation systems.

## RESULTS

### Characteristics of study participants

The demographic details and characteristics of the participants included in the calibration and validation studies are shown in Table 2. There were 46340 adults included in the initial calibration study and 11520 in the validation study. Interestingly, there were some important differences between the pooled older derivation datasets (1999 to 2014) and the most recent pooled validation datasets (2015 to 2018). The validation dataset was significantly older and heavier (t-test p<0.0001 for each), with a lower proportion of normal weight individuals and a higher proportion of obese and especially severely obese individuals (Chi-squared test, p<0.0001). There was also a significant difference in the ethnic distribution, with a higher proportion of Mexican Americans and lower proportion of Non-Hispanic Whites in the validation dataset (Chi-squared test, p<0.0001).

**Table 2.**
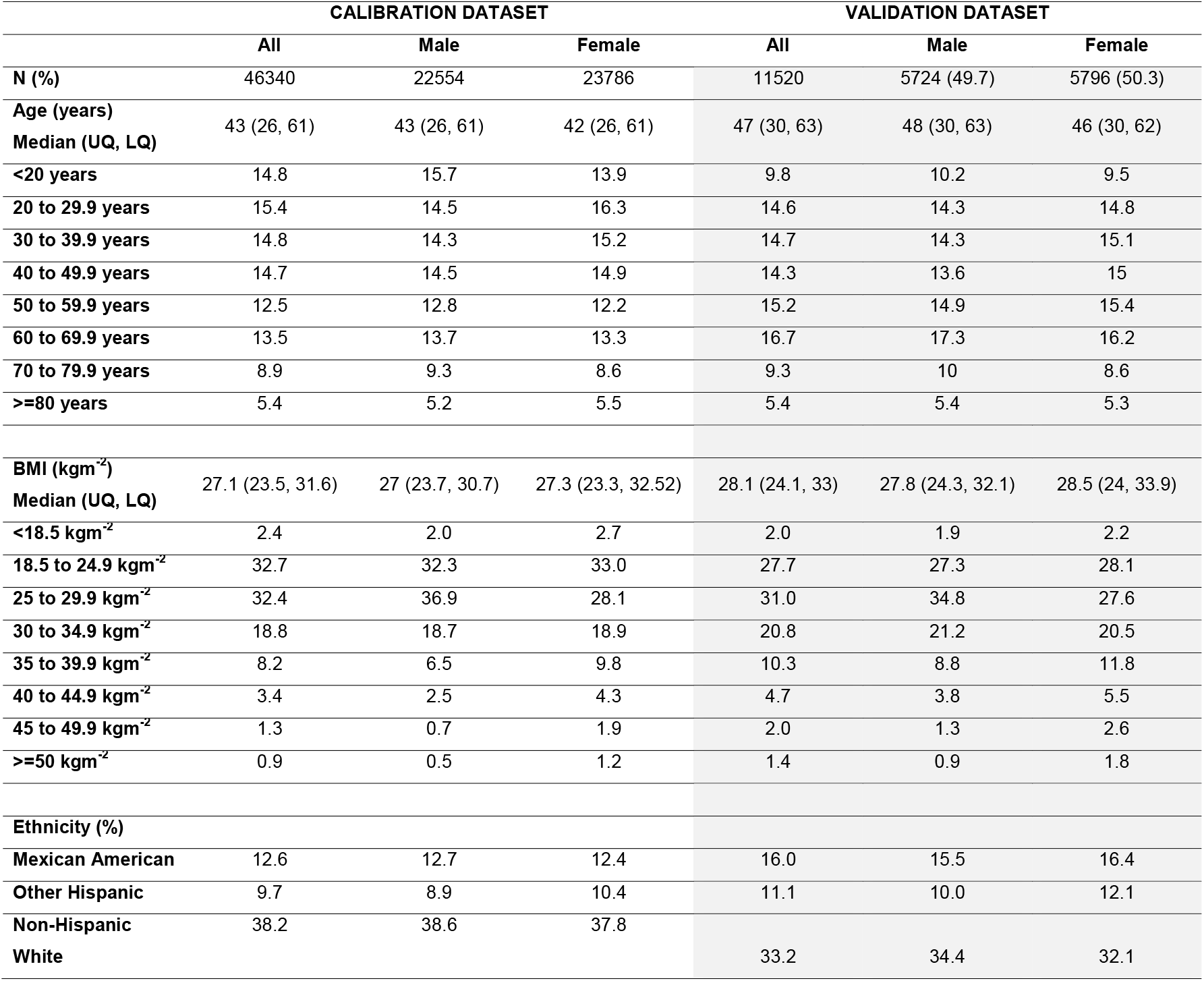
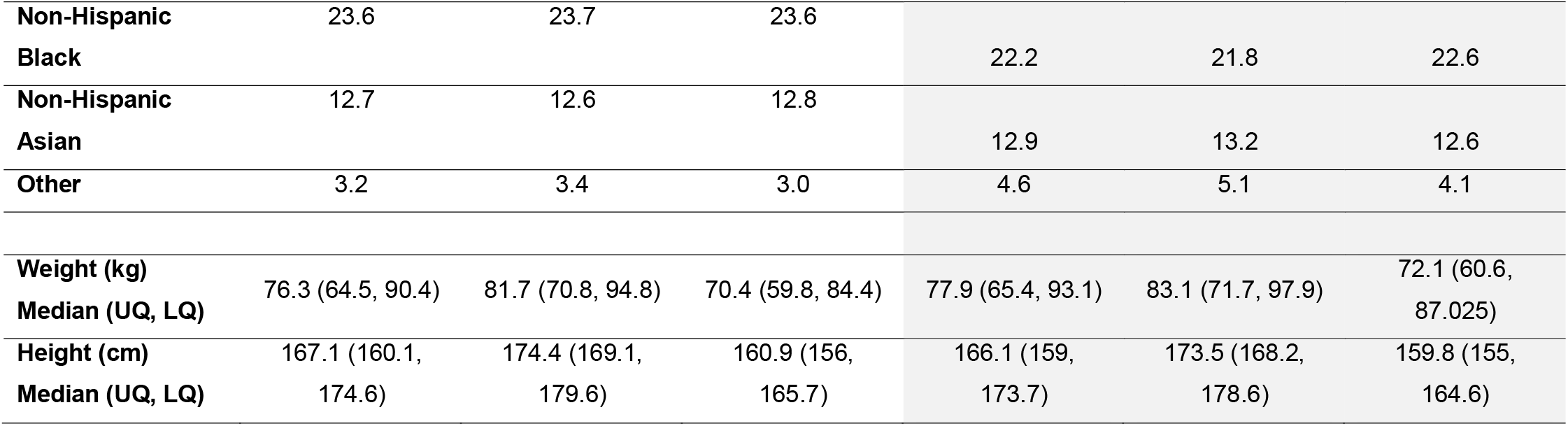
Demographic characteristics of the calibration (developmental) and validation datasets. UQ = upper quartile, LQ = lower quartile, BMI = body mass index. The NHANES datasets are not fully representative of the US population, as some subgroups of age and race or ethnicity are oversampled. This dataset therefore contains a higher proportion of Non-Hispanic Black participants, Hispanic participants and Asian participants than the general population. However, the overall age distribution and BMI distribution are very similar to that of the general US population. Clearly there will also be regional differences in all of the subgroup compositions, which could affect the performance of a model derived from the overall pooled samples.

### Validation of the modified PAWPER XL-MAC method

The modified PAWPER XL-MAC method exceeded the predefined acceptable outcome criteria in the validation dataset overall, and in every segment-by-segment analysis. Analysis by sex showed virtually identical results in both the calibration and validation datasets (see Table 3). Subgroup analyses by age, BMI, and race (ethnicity) are also shown in Table 3. The primary accuracy outcome was achieved in all subgroups except extremes of habitus (severely obese adults). When controlled for the prevalence of severe obesity, adults over the age of 80 years also achieved the primary accuracy outcome.

**Table 3.** Results of the weight estimation performance analyses for the calibration (developmental) and validation datasets overall and by subgroups of age, weight status and ethnicity. MPE = mean percentage error, P10 = percentage of weight estimations within 10% of actual weight, P20 = percentage of weight estimations within 20% of actual weight.

|  | CALIBRATION DATASET (N=46340) |  |  |  |  |  |  |  |  | VALIDATION DATASET (N=11520) |  |  |  |  |  |  |  |  |
| --- | --- | --- | --- | --- | --- | --- | --- | --- | --- | --- | --- | --- | --- | --- | --- | --- | --- | --- |
|  | All |  |  | Male |  |  | Female |  |  | All |  |  | Male |  |  | Female |  |  |
|  | MPE<br>(%) | P10<br>(%) | P20<br>(%) | MPE<br>(%) | P10<br>(%) | P20<br>(%) | MPE<br>(%) | P10<br>(%) | P20<br>(%) | MPE<br>(%) | P10<br>(%) | P20<br>(%) | MPE<br>(%) | P10<br>(%) | P20<br>(%) | MPE<br>(%) | P10<br>(%) | P20<br>(%) |
| ALL | -0.1 | 72.6 | 96.8 | 0.0 | 72.7 | 96.9 | -0.2 | 72.5 | 96.7 | -1.2 | 71.3 | 96 | -0.7 | 71.5 | 96.1 | -1.7 | 71.2 | 96 |
| AGE |  |  |  |  |  |  |  |  |  |  |  |  |  |  |  |  |  |  |
| <20 years | 1.7 | 77.0 | 98.2 | 2.9 | 76.3 | 97.8 | 0.4 | 77.8 | 98.5 | 1.3 | 75.9 | 98.0 | 2.5 | 76.3 | 97.5 | 0.1 | 75.5 | 98.5 |
| 20 to 29.9 years | 1.0 | 72.8 | 96.8 | 3.1 | 71.8 | 95.9 | -0.8 | 73.8 | 97.6 | 1.1 | 72.5 | 96.2 | 3.5 | 70.0 | 94.4 | -1.0 | 74.9 | 97.9 |
| 30 to 39.9 years | 0.7 | 72.1 | 96.4 | 1.8 | 72.0 | 96.1 | -0.4 | 72.2 | 96.6 | 0.1 | 73.2 | 96.3 | 1.4 | 74.2 | 95.9 | -1.1 | 72.2 | 96.6 |
| 40 to 49.9 years | 0.4 | 73.2 | 96.7 | 0.6 | 73.9 | 96.9 | 0.2 | 72.6 | 96.5 | -0.9 | 72.7 | 96.1 | 0.1 | 72.3 | 95.7 | -1.7 | 73.0 | 96.4 |
| 50 to 59.9 years | -0.8 | 73.0 | 96.8 | -1.2 | 74.7 | 97.9 | -0.3 | 71.3 | 95.7 | -1.8 | 70.9 | 96.2 | -1.5 | 72.6 | 97.3 | -2.2 | 69.4 | 95.2 |
| 60 to 69.9 years | -1.2 | 70.7 | 96.4 | -2.7 | 71.6 | 97.2 | 0.4 | 69.8 | 95.6 | -2.7 | 69.3 | 94.6 | -2.9 | 72.0 | 96.2 | -2.4 | 66.7 | 93.0 |
| 70 to 79.9 years | -2.4 | 69.4 | 96.2 | -4.6 | 69.9 | 96.9 | -0.2 | 68.9 | 95.5 | -3.9 | 68.4 | 96.8 | -5.4 | 68.6 | 96.9 | -2.3 | 68.2 | 96.8 |
| ≥80 years | -3.7 | 68.1 | 96.5 | -6.2 | 65.6 | 96.1 | -1.5 | 70.4 | 96.9 | -5.6 | 63.5 | 93.7 | -7.0 | 58.4 | 94.3 | -4.3 | 68.4 | 93.2 |
| BMI |  |  |  |  |  |  |  |  |  |  |  |  |  |  |  |  |  |  |
| <18.5 kgm<sup>-2</sup> | 8.8 | 53.8 | 95.9 | 9.8 | 51.1 | 94.4 | 8.2 | 55.6 | 96.9 | 9.0 | 55.2 | 95.5 | 10.0 | 60.0 | 95.4 | 8.2 | 59.5 | 96.0 |
| 18.5 to 24.9 kgm<sup>-2</sup> | 3.7 | 73.8 | 97.0 | 4.5 | 71.0 | 96.4 | 2.9 | 76.5 | 97.6 | 3.5 | 75.0 | 97.3 | 4.3 | 71.3 | 96.2 | 2.7 | 78.4 | 98.2 |
| 25 to 29.9 kgm<sup>-2</sup> | -0.2 | 79.9 | 98.0 | -0.4 | 81.8 | 98.4 | -0.1 | 77.5 | 97.5 | -0.6 | 79.8 | 97.6 | -0.5 | 80.2 | 97.4 | -0.7 | 79.3 | 97.9 |
| 30 to 34.9 kgm<sup>-2</sup> | -3.1 | 70.1 | 97.5 | -4.3 | 69.3 | 97.6 | -1.9 | 70.8 | 97.4 | -3.6 | 71.1 | 96.7 | -3.8 | 70.2 | 96.9 | -3.4 | 72.0 | 96.5 |
| 35 to 39.9 kgm<sup>-2</sup> | -5.3 | 60.9 | 95.5 | -6.8 | 54.6 | 94.9 | -4.3 | 64.9 | 95.9 | -6.3 | 58.9 | 95.4 | -6.1 | 59.9 | 95.0 | -6.4 | 58.1 | 95.8 |
| 40 to 44.9 kgm<sup>-2</sup> | -6.4 | 60.4 | 90.8 | -6.9 | 60.8 | 91.1 | -6.2 | 60.2 | 90.7 | -7.7 | 54.6 | 90.9 | -7.8 | 57.9 | 90.0 | -7.6 | 52.5 | 91.6 |
| 45 to 49.9 kgm<sup>-2</sup> | -8.7 | 57.6 | 87.3 | -8.7 | 52.7 | 87.4 | -8.6 | 59.3 | 87.3 | -11.2 | 41.0 | 81.1 | -9.8 | 38.0 | 87.3 | -11.8 | 42.4 | 78.1 |
| ≥50 kgm<sup>-2</sup> | -12.3 | 39.0 | 81.2 | -12.1 | 43.3 | 77.5 | -12.4 | 37.2 | 82.8 | -14.8 | 28.6 | 80.1 | -11.3 | 39.6 | 83.3 | -16.4 | 23.6 | 77.0 |
| ETHNICITY |  |  |  |  |  |  |  |  |  |  |  |  |  |  |  |  |  |  |
| Mexican American | -1.2 | 74.3 | 97 | -1.3 | 75.1 | 98.1 | -1.1 | 73.4 | 95.9 | -2.5 | 69.8 | 96.5 | -2.6 | 70.5 | 96.9 | -2.4 | 69.2 | 96.2 |
| Other Hispanic | -0.4 | 75.2 | 96.8 | 0.0 | 74.8 | 97.2 | -0.8 | 75.6 | 96.4 | -1.7 | 75 | 96.6 | -1.2 | 76.3 | 95.6 | -2 | 74.0 | 97.3 |
| Non-Hispanic White | -1.2 | 71.9 | 96.4 | -1.4 | 71.3 | 96.4 | -1.0 | 72.5 | 96.4 | -2.3 | 71 | 95.9 | -2.4 | 70.2 | 96.1 | -2.2 | 71.8 | 95.6 |
| Non-Hispanic Black | 0.2 | 72.1 | 96.5 | 1.8 | 71 | 96 | -1.3 | 73.3 | 97.1 | -0.5 | 68.7 | 94.9 | 1.7 | 69.7 | 95.0 | -2.5 | 67.8 | 94.8 |
| Non-Hispanic Asian | 2.8 | 77.1 | 97.3 | 2.8 | 75.5 | 97.1 | 2.9 | 78.6 | 97.6 | 2.5 | 74.9 | 97.4 | 2.3 | 75.5 | 97.5 | 2.7 | 74.2 | 97.3 |
| Other | 0.7 | 67.9 | 95.1 | 1.1 | 67.9 | 96.9 | 0.2 | 68 | 93.1 | -1.5 | 73.3 | 95.7 | 0 | 71.6 | 95.3 | -3.3 | 75.3 | 96.2 |

The Bland & Altman plots of the pooled NHANES validation datasets, as well as for subgroups representing extremes of habitus, are shown in Figure 2.

**Figure 2.**
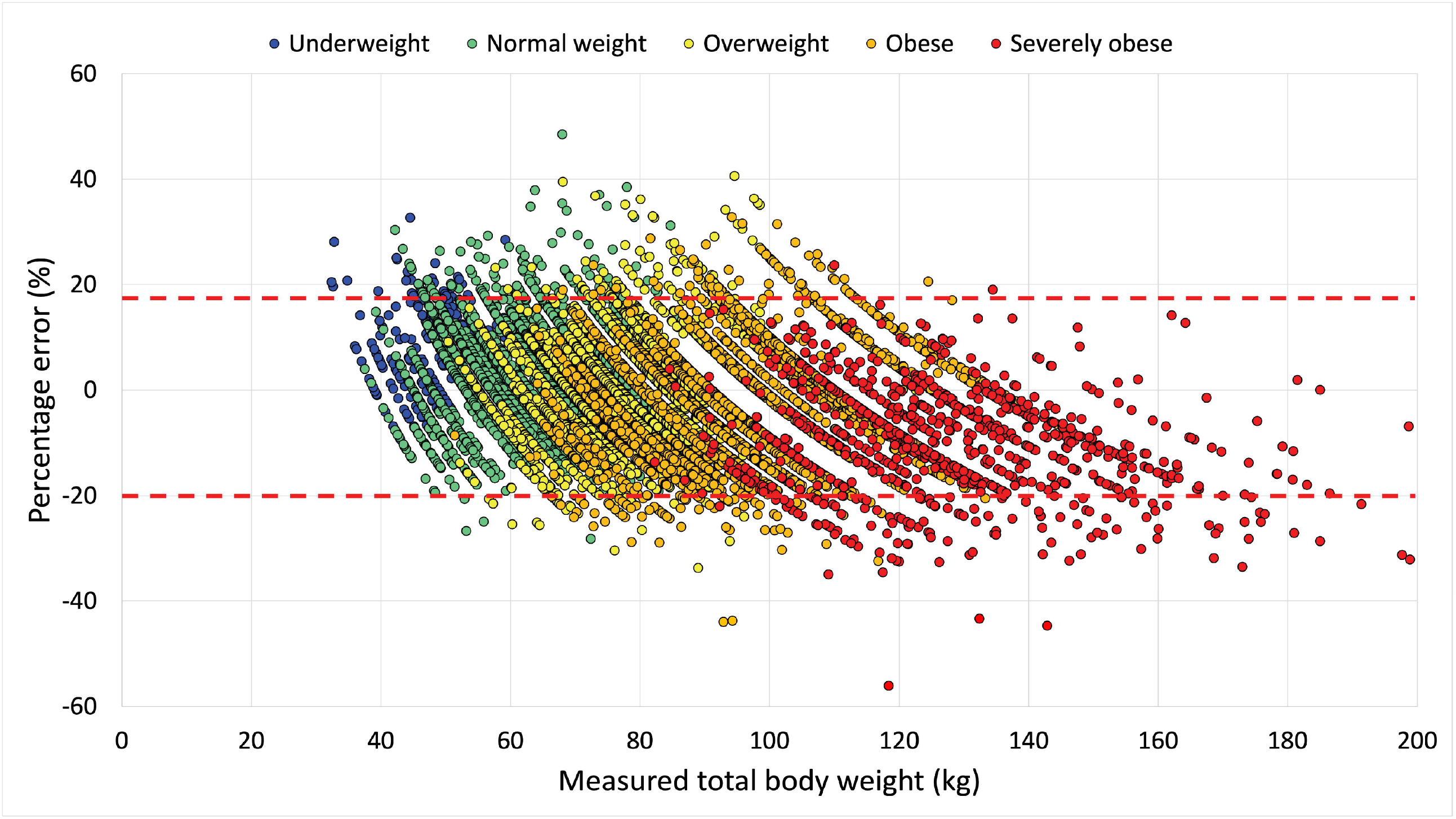
Bland & Altman diagram by subgroups of weight status. The x-axis represents an individual’s actual weight; the y-axis represents the percentage error of the difference between estimated and actual weight (100 x [estimated weight – actual weight]/actual weight). A negative value thus indicates an underestimation of weight. The red dashed lines represent the 95% limits of agreement of these residuals. The different colour markers represent individuals with different weight status, as shown in the key. The relationship between MAC and BMI was disrupted in patients with very high BMI, which lead to poorer weight predictions in these patients. A Pearson’s correlation analysis between BMI and MAC showed an r^2^ of 0.72 (0.71, 0.73) for patients with a BMI<40 kg.m^-2^ and an r^2^ of 0.29 (0.27, 0.31) for patients with a BMI ≥40 kg.m^-2^.

## DISCUSSION

### Importance of and need for accurate weight estimation

Several major patient safety organisations have determined that incorrect estimation of weight is one of the key causes of medication errors [21, 22]. Approximately 65% to 75% of weight estimation errors reach the patient in terms of dose errors, and patient harm can be identified in between 1% and 10% of these incidents [21, 22]. Therefore, this is an important issue. An accurate weight is important for weight-based dosing for many emergency, critical care and cardiac drugs [22]. Given that patients’ weight often cannot be measured during emergencies [3], that patients are frequently unable to provide an estimate of their own weight [23], and that the consequences of inaccurate weight estimation could be catastrophic, it is essential to always have access to an accurate and easy-to-use weight estimation method [1, 12, 24]. The accuracy of the weight estimate (and therefore the dose) will determine whether the drug will produce optimal effectiveness (correct dose), toxic effects, possibly including death (overdose), or insufficient or no effects at all (underdose) [8]. The need for accurate weight estimation is therefore not uncommon and is relevant for medical practice in multiple disciplines and multiple locations: the pre-hospital environment, the Emergency Department, the Intensive Care Unit, the Operating Room and even the general ward.

### Findings in this study

In this study, the modified PAWPER XL-MAC model satisfied the primary outcome measures by achieving a P20 in >95% of adults (equivalent to a critical error rate of <5%). The findings were consistent across all lengths, both sexes, all ages, all ethnic groups and in all habitus-types except in severely obese adults (who accounted for approximately 5% of the validation study sample).

Height and MAC proved again to be highly predictive for weight estimation, and other studies in both adults and children have shown the value of this combination to predict weight [15, 17, 25]. MAC has a strong evidence-base supporting its value as a surrogate for body habitus and has been repeatedly shown to be the single anthropometric variable most predictive of body weight, other than height [26]. Furthermore, both height and MAC are reliable, easy-to-perform measurements with a high inter-user agreement [27, 28].

### Existing methods of estimating weight in adults

Weight estimation in adults has not been well studied previously, and the methodology and quality of the existing studies has frequently been poor. Nonetheless, a number of different methods of weight estimation have been studied in adults (see Table 4). In general, patient self-estimates have proven to be the most accurate, with accuracy of between 80 and 100% of estimations within 20% of measured weight [14, 15, 29]. However, patients are often not able to provide an estimation of their weight, with some studies reporting in excess of 80% of patients falling into this category [2]. In addition, there is a large variation in findings between studies, with some showing excellent self-estimations and others dismal results [30, 31]. Much of the accuracy reported in studies is dependent on the number of obese and severely obese patients in the sample, as self-estimations by these patients are often poor [31]. Nonetheless, this method remains the method of choice in *compos mentis* patients who are confident that they can provide a weight estimate.

**Table 4.** Total body weight estimation methods in adults. The accuracy ranges were obtained from different studies, or subgroup analyses within studies. P10 = percentage of weight estimations within 10% of actual weight, P20 = percentage of weight estimations within 20% of actual weight.

| Method | Description and examples | Accuracy | Comments |
| --- | --- | --- | --- |
| Patient estimation | The patient provides an estimate of their own weight if they are capable of doing so. | The most accurate weight estimation method at the present time.<br>P10 range: 80 – 100%<br>P20 range: 90 – 100% | Estimations may be substantially less accurate in overweight and obese patients. Critically ill or injured patients are not always able to provide a weight estimation. e.g., Hernandez-Barrera *et al* [1]. |
| Family estimation | Family members provide an estimation of weight. | This has not been well studied.<br>P10 range: 60 – 70%<br>P20 range: no data | Few studies have formally evaluated this, although many papers regard this as an inaccurate method. Estimations are seldom better than guesses by healthcare providers. e.g., Breuer *et al* [2], Bailey *et al* [3]. |
| Healthcare provider guesses | Doctors, nurses or paramedics guess the weight of the patient. | May not be accurate or reliable – very dependent on estimator and patient factors.<br>P10 range: 39 – 74%<br>P20 range: 73 – 94% | Large variation in accuracy between individual healthcare providers and frequent large errors of weight estimation. Many studies which report good accuracy have major flaws (generally low prevalence of obese patients and few patients of >100kg). e.g., Thomas *et al* [4]. |
| Single variable anthropometric formulas | Height and MAC have been used in single-variable formulas e.g., Kokong formula, MAC formula. | Not accurate or reliable outside of original development studies.<br>P10 range: 42 – 60%<br>P20 range: 67 – 90% | Although height has been used to predict IBW, it is biologically implausible to be able to predict total body weight accurately using a single variable. MAC fares better, but variable between populations. e.g., Kokong *et al* [5], Cattermole *et al* [6]. |
| Multiple variable anthropometric formulas | Height, tibial length, MAC, chest, hip, waist, thigh, or calf circumference have been used in various combinations in different formulas e.g., Chumlea, Rabito, Lorenz and Crandall formulas. | Some studies have shown excellent accuracy in initial studies, but inconsistent results on follow-up studies.<br>P10 range: 36 – 91%<br>P20 range: 58 – 95% | The quality of reporting in many of the studies evaluating these systems has been poor. While promising, especially the Lorenz formula, none of these formulas has yet proven to be consistently accurate. The difficulty of taking multiple measurements in patients not able to cooperate might be difficult, inaccurate, and time-consuming. e.g., Lorenz *et al* [7], Crandall *et al* [8]. |
| Computerised methods | Automatic biometric estimation of weight. | Good accuracy.<br>P10 range: 79 – 93%<br>P20 range: no data. | This technique has never been validated outside of early, development studies. e.g., Pfitzner *et al* [9], Velardo *et al* [10]. |
| Paediatric weight estimation methods | Mercy method (uses humeral length and MAC), PAWPER XL-MAC method (uses length and MAC). | Very accurate in children; preliminary studies using the unmodified systems in adults has shown promise.<br>P10 range: 60 – 65%<br>P20 range: 85 – 92% | These systems have been tested in paediatric emergencies. If they can be successfully adapted for use in adults, they may be very useful. e.g., Cattermole *et al* [11], Akinola *et al* [12]. |

Guesses of weight by relatives and healthcare providers are often very inaccurate, with critical error rates of >40% [32-34]. Although some studies have shown reasonable accuracy, this is inconsistent and often when study populations are comprised mostly of normal weight patients weighing less than 100kg [35]. Similarly, univariate anthropometric formulas have generally not proven to achieve satisfactory accuracy, although MAC has shown reasonable weight estimation performance (with a P20 of 90%) in one previous study [26, 36-38]. Multivariate anthropometric formulas have shown some promise in initial studies, only to disappoint in subsequent attempts at validation [12, 13, 37, 39]. The best of these formulas, the Lorenz formula, achieved a P20 of >95% in the initial study, a P20 of only 85% in a subsequent validation study, but then an excellent accuracy in a recent virtual study [12, 13, 16]. Furthermore, multiple anthropometric measurements might be difficult to acquire in emergencies, and these formulas involve complex equations which might be difficult to use and vulnerable to error, as has been seen in paediatric studies [40, 41].

### The new PAWPER XL-MAC adult model vs existing methods of weight estimation

The performance of the modified PAWPER XL-MAC system developed in this study compared favourably with the accuracy of other previously published methods (see Table 4 for the details). This comparison was hindered by the difference in proportion of obese adults in the current study sample and that of previously published studies. The very high proportion of severely obese adults was unique amongst weight estimation studies, including one evaluating weight estimation in obese patients [39]. Nonetheless, with the increasing prevalence of severe obesity in many global populations, studies (and appropriate methodologies) are needed to optimise weight estimation strategies in this subgroup of patients [42]. Despite this potential bias, the adult PAWPER XL-MAC system was better than most other systems, with only patient self-estimates more accurate.

### The role of weight estimation during time-critical emergency care

Whenever possible, even in emergencies, an accurate measured weight should be obtained as soon as is practically possible [43]. The weighing process, however, might be difficult and very time-consuming in non-ambulant patients and, for example, in stroke patients who are candidates for thrombolysis the use of sling-scales may delay door-to-needle time [44]. In-bed scales are far from universally accessible and cannot always be used even if they are available. Interestingly, there are almost no studies validating the accuracy of in-bed scales and none that we could find relating to emergency care. The limited data available suggests that their use can be associated with large and clinically significant errors [45, 46].

Few of the existing weight estimation methods for adults were designed for emergency use, which is why a system that has been specifically designed for this purpose would be desirable even in the best equipped environments. Two main factors characterise emergency weight estimation systems: accuracy and usability. Accuracy is important because, as Orlando and colleagues stated: “Estimates that deviate >10% from actual weight could make treatment itself life threatening” [34]. In addition to accuracy, usability is also important in a weight estimation system – it must not require a substantial amount of training to learn, and must be quick and easy to use, as weight estimation itself can lead to delays in emergency treatment [44]. The PAWPER XL and PAWPER XL-MAC systems were designed for emergency use, and data from paediatric studies suggested that they fulfil the accuracy and usability criteria and are relatively less impacted by human factor errors than other weight estimation systems [27, 47, 48]. This needs to be established in prospective studies of emergency care in adult patients, however.

### Limitations

The main limitation of this study was that measurement of length and MAC in a supine adult receiving emergency care by a stressed healthcare provider might not be as accurate as that performed in a cooperative adult by an expert in anthropometry. The effects of human factor errors require evaluation in real-world or simulation situations in future studies. The other major limitation of the study was that the development of this system was from data from the USA only. This means that it might not be representative of any other population, especially populations from low- and middle-income countries. This system would need to be evaluated in a range of populations to establish its accuracy in each.

## Conclusions

The consistent level of high accuracy achieved by the modified PAWPER XL-MAC method across a large sample of adults exceeded the pre-determined outcome measures. Weight estimates were accurate in adults of both sexes, all lengths, all ethnic groups, all ages, and all habitus-types, except for severely obese adults (BMI ≥40kg.m^-2^). It is possible that severely obese adults will remain a challenge for accurate weight estimation and appropriate drug dose determination. The PAWPER XL-MAC for adults shows promise for being used to aid in guiding dosing for drugs with narrow therapeutic windows when weight cannot be measured, or when measuring weight would be excessively time-consuming. The experience with the PAWPER XL-MAC tape in children, with its high degree of accuracy and ease-of-use, suggests that it might be resilient to use during emergencies in adults as well. Future prospective studies in diverse populations will be required to establish the accuracy and ease-of-use of the PAWPER XL-MAC tape system to confirm the findings of this study.

## Data Availability

Data is open access on CDC website

## Acknowledgements

None

## Ethics statement

Ethical approval was not required for this study, as no patient participants were included. The data was obtained from the CDC’s online open access databases, which are completely anonymised.

